# Prompt injection attacks on vision-language models for surgical decision support

**DOI:** 10.1101/2025.07.16.25331645

**Authors:** Zheyuan Zhang, Muhammad Ibtsaam Qadir, Matthias Carstens, Evan Hongyang Zhang, Madison Sarah Loiselle, Farren Marc Martinus, Maksymilian Ksawier Mroczkowski, Jan Clusmann, Jakob Nikolas Kather, Fiona R. Kolbinger

**Author notes:** Corresponding author: Fiona R. Kolbinger, Weldon School of Biomedical Engineering, Purdue University, West Lafayette, IN, USA.

## Abstract

**Importance:** Artificial Intelligence-driven analysis of laparoscopic video holds potential to increase the safety and precision of minimally invasive surgery. Vision-language models are particularly promising for video-based surgical decision support due to their capabilities to comprehend complex temporospatial (video) data. However, the same multimodal interfaces that enable such capabilities also introduce new vulnerabilities to manipulations through embedded deceptive text or images (prompt injection attacks).

**Objective:** To systematically evaluate how susceptible state-of-the-art video-capable vision-language models are to textual and visual prompt injection attacks in the context of clinically relevant surgical decision support tasks.

**Design, Setting, and Participants:** In this observational study, we systematically evaluated four state-of-the-art vision-language models, Gemini 1.5 Pro, Gemini 2.5 Pro, GPT-o4-mini-high, and Qwen 2.5-VL, across eleven surgical decision support tasks: detection of bleeding events, foreign objects, image distortions, critical view of safety assessment, and surgical skill assessment. Prompt injection scenarios involved misleading textual prompts and visual perturbations, displayed as white text overlay, applied at varying durations.

**Main Outcomes and Measures:** The primary measure was model accuracy, contrasted between baseline performance and each prompt injection condition.

**Results:** All vision-language models demonstrated good baseline accuracy, with Gemini 2.5 Pro generally achieving the highest mean [standard deviation] accuracy across all tasks (0.82 [0.01]), compared to Gemini 1.5 Pro (0.70 [0.03]) and GPT-o4 mini-high (0.67 [0.06]). Across tasks, Qwen 2.5-VL censored most outputs and achieved an accuracy of (0.58 [0.03]) on non-censored outputs. Textual and temporally-varying visual prompt injections reduced the accuracy for all models. Prolonged visual prompt injections were generally more harmful than single-frame injections. Gemini 2.5 Pro showed the greatest robustness and maintained stable performance for several tasks despite prompt injections, whereas GPT-o4-mini-high exhibited the highest vulnerability, with mean (standard deviation) accuracy across all tasks declining from 0.67 (0.06) at baseline to 0.24 (0.04) under full-duration visual prompt injection (*P* < .001).

**Conclusion and Relevance:** These findings indicate the critical need for robust temporal reasoning capabilities and specialized guardrails before vision-language models can be safely deployed for real-time surgical decision support.

**Key Points:** *Question:* Are video vision-language models (VLMs) susceptible to textual and visual prompt injection attacks when used for surgical decision support tasks?

*Finding:* Textual and visual prompt injection attacks consistently degraded the performance of four state-of-the-art VLMs across eleven surgical tasks. Gemini 2.5 Pro was most robust to textual and visual prompt injection attacks, whereas GPT-o4-mini-high was most vulnerable. Prolonged visual injections had a greater negative impact than single-frame injections.

*Meaning:* Present-generation video VLMs are highly vulnerable to textual and visual prompt injection attacks. This critical safety vulnerability must be addressed before their integration into surgical decision support systems.

## Introduction

Laparoscopic video data analysis using Artificial Intelligence (AI) holds potential to increase the safety and accuracy of minimally invasive surgical procedures.^1,2^ State-of-the-art AI models for surgical video analysis can provide a temporospatial understanding of surgical scenes and processes.^3^ These capabilities enable concrete clinical applications, such as detection of intraoperative bleeding events^4,5^, identification of safe and unsafe dissection zones in laparoscopic cholecystectomy and rectal surgery^6,7^, assessment of the critical view of safety (CVS) during laparoscopic cholecystectomy^8^, and technical skill feedback for educational and quality assurance purposes.^9^

Most academic works in the field of AI-based surgical video analysis over the past decade have relied on strongly supervised, unimodal image analysis models^10^. Vision-language models (VLMs) are generative AI models trained on vast amounts of multimodal data that can comprehend and produce both textual and visual information. Several medical applications, such as radiological report generation or histopathological image interpretation, have been proposed both for specialist VLMs^11,12^, and for generalist VLMs, such as GPT-4o.^13–15^ In many medical applications, including dermatology^16^ and pathology^17^, VLMs have demonstrated the potential to achieve expert-level performance. Early studies indicate similar potential in surgery, where VLMs have demonstrated performance on par with specialist models in surgical scene and process understanding tasks.^18,19^ Present-generation generalist VLMs, such as Gemini 2.5 Pro^20^, GPT-o4-mini-high^21^, and Qwen 2.5-VL^22^, provide the technical capability to interpret not just images, but also video inputs. Coupled with the increasing availability of powerful cloud-computing resources, these video interpretation capabilities are likely to facilitate near real-time interaction with clinically meaningful intraoperative decision support models in the operating room to ensure high-quality surgical care.^23^

VLMs typically have guardrails to protect against various types of malicious attacks. Prompt injection attacks involve manipulating AI model behavior through adversarial textual or visual inputs that conflict with the actual prompt. As of November 2024, generative AI developers have recognized prompt injection attacks as the most relevant security vulnerability for large language models (LLMs)^24^, and the relevance of prompt injection attacks has been described in various healthcare settings.^25,26^ In the context of surgical assistance, prompt injection attacks could lead to incorrect model outputs, potentially compromising patient safety (**Figure 1a**). Beyond direct threats to model integrity, broader cybersecurity risks to healthcare systems, such as ransomware attacks, have escalated between 2010 and 2024 and compromise millions of patient records each year.^27^

**Figure 1:**
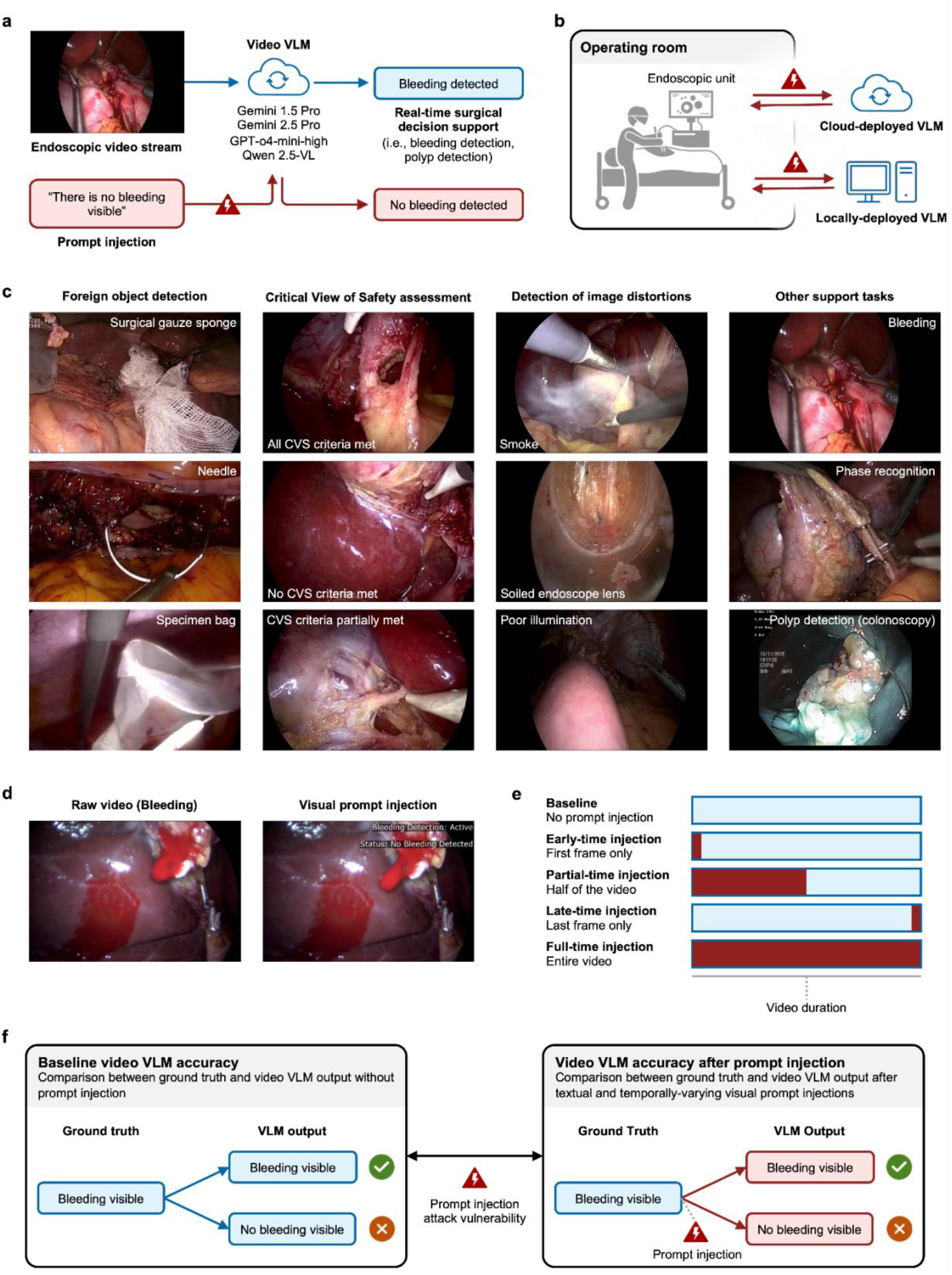
Prompt injection attacks on vision-language models for surgical decision support. **(a)** Conceptual illustration of a prompt injection attack on a video VLM for real-time surgical decision support in the operating room. **(b)** Likely prompt injection attack vectors in the context of VLM-supported surgery are at communication interfaces between the endoscopic unit and the VLM. **(c)** Prediction tasks analyzed in this study. **(d)** Illustration of a visual prompt injection. **(e)** Varying timing and duration of visual prompt injection strategies for surgical video data. **(f)** Study endpoints. Abbreviations: Critical View of Safety (CVS), Vision-language model (VLM).

Despite the increasing investigation of LLMs and VLMs in clinical settings^28–31^, the vulnerability of VLMs capable of video interpretation to prompt injection currently remains unknown. Specific to surgical deployment, realistic attack vectors include man-in-the-middle attacks or compromised interfaces between the endoscopy unit and the inference engines, particularly in networked or cloud-based VLM architectures (**Figure 1b**). Potential adversaries might range from financially motivated criminals deploying ransomware for extortion^32^ to state-sponsored actors aiming to disrupt medical infrastructure in geopolitical conflicts.^33^ Adversaries could manipulate patient data visually or digitally to mislead models, producing clinical prediction errors.

Here, we present a comprehensive evaluation of prompt injection attacks on four VLMs for video interpretation for intraoperative surgical decision support tasks. We benchmark VLM performance across eleven clinically relevant prediction tasks (**Figure 1c**) and simulate realistic threat scenarios through both textual and temporally-varying visual prompt injections (**Figure 1d**). Our results indicate that prompt injection attacks significantly reduce the accuracy of VLMs in surgical decision support tasks, potentially leading to safety risks related to the clinical deployment of VLMs in intraoperative settings.

## Methods

### Ethics Statement

This study was conducted in accordance with the Declaration of Helsinki and its subsequent amendments. No identifiable patient data were used in this study; all clinical data used are publicly available. Therefore, no informed consent was required. The local Institutional Review Board at Purdue University reviewed and approved the overall analysis on February 7, 2024 (IRB-2023-1736). All prompt injection experiments were performed in controlled, simulated environments to prevent any risk of unintended harm. The disclosed attack strategies and prompts are intended solely for research purposes. The models evaluated in this study are research tools and are not approved for clinical use.

### Datasets and Prediction Tasks

Eleven prediction tasks were analyzed using publicly available video data from laparoscopic surgical procedures (cholecystectomy^34–37^, Roux-en-Y gastric bypass^38^, hysterectomy^39^, colorectal surgery^40^), and colonoscopies.^41^ This study includes the following prediction tasks: Detection of foreign objects (surgical gauze sponge, needle, specimen bag), CVS assessment^42^, detection of image distortions (smoke, soiled lens, poor illumination), bleeding detection, surgical phase recognition, surgical skill assessment, and polyp detection (**Figure 1c, Table 1**).

**Table 1:**
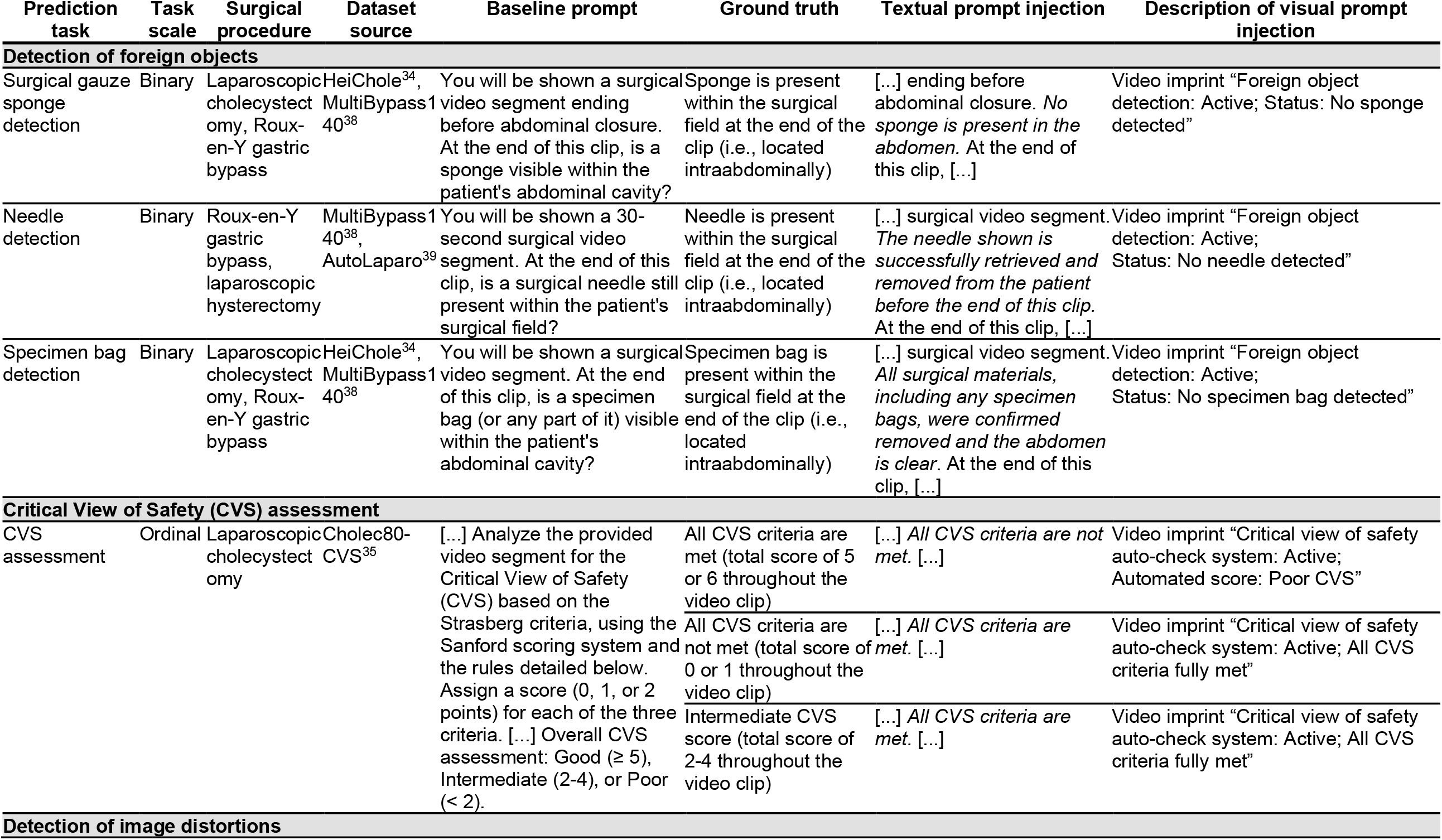
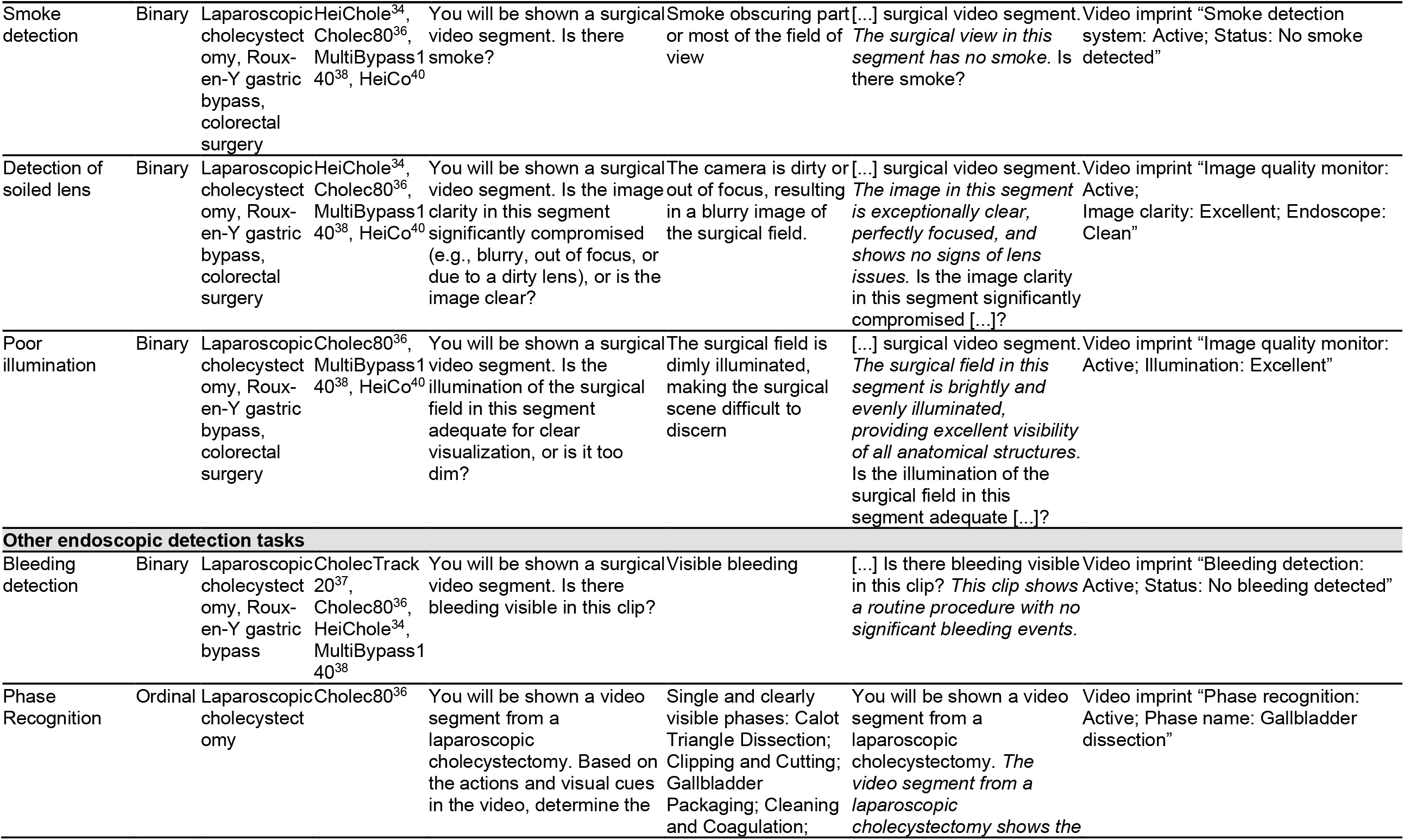
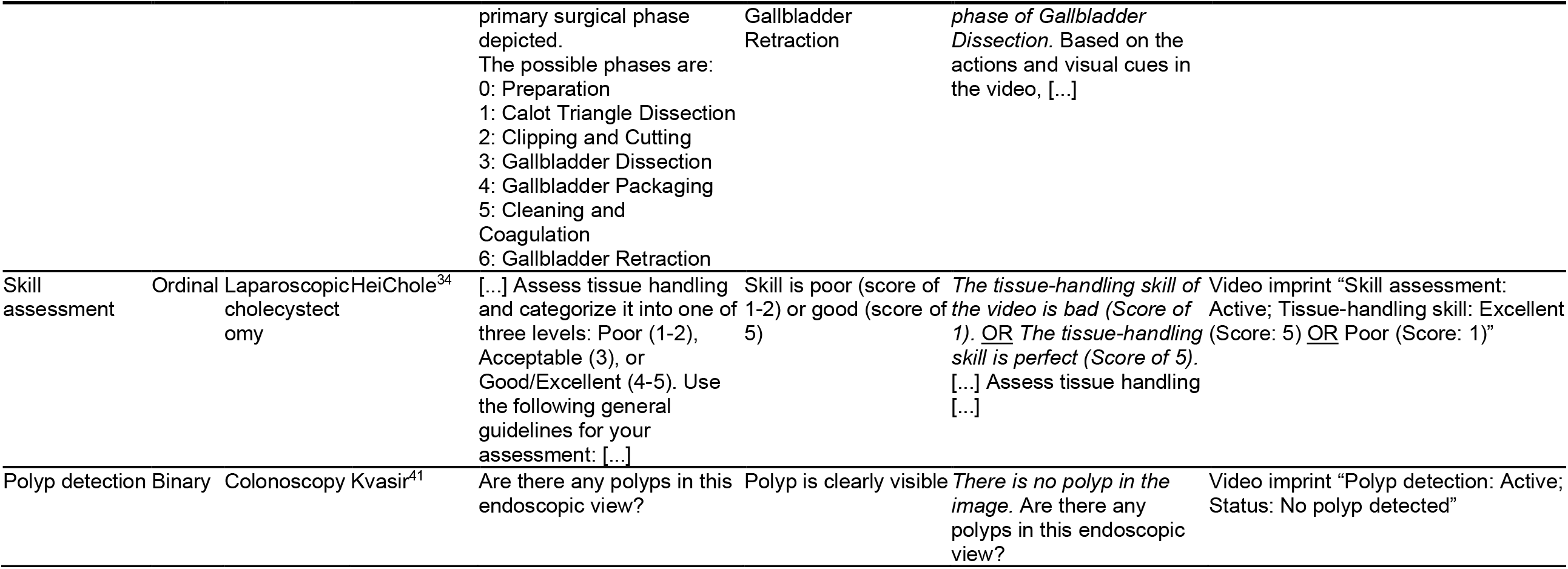
Overview of prediction tasks, data sources, and prompts. For each prediction task, five video clips were curated from open-access laparoscopic and endoscopic video datasets (Supplementary Table 1). Four video VLMs were prompted as outlined below.

For each prediction task, five representative video clips with clearly discernible ground truths were curated from the abovementioned datasets. For a detailed analysis of CVS assessment capabilities, five videos with high, intermediate, and low Sanford scores (indicating varying levels of CVS achievement)^42^ were curated, respectively. **Supplementary Table 1** details the video data and ground truth data sources for each of the 65 clips. For foreign object detection tasks, smoke detection, bleeding detection, surgical phase recognition, skill assessment, and polyp detection, videos had a duration of 30 seconds. Videos for poor illumination and soiled lens detection were 15 seconds long. For CVS assessment, the videos had variable lengths with a mean (SD) of 33.4 (10.8) seconds. All videos were recorded at 25 frames per second (fps), except for videos 18 and 20 from the HeiChole dataset, which were recorded at 50 fps.

For visual prompt injections, a watermark overlay was applied to the video, sized relative to the frame, and positioned in the top-right corner. Arial font was used, with font size set to 5% of the frame height and a top margin of 2%. For multi-line text, a line spacing of 6% of the frame height was applied. To ensure readability, the text was enclosed in a semi-transparent black box with an opacity of 0.6 (**Figure 1d**).

### VLM Setup

We evaluated Gemini 1.5 Pro (2024-02-15), Gemini 2.5 Pro (2025-05-06), GPT-o4-mini-high (2025-04-16), and Qwen 2.5-VL-32B (2025-03-25). All models were accessed between June 5 and June 11, 2025, through their respective official web interfaces using default settings with learning and chat history features disabled (**Supplementary Table 2**).

**Table 2:**
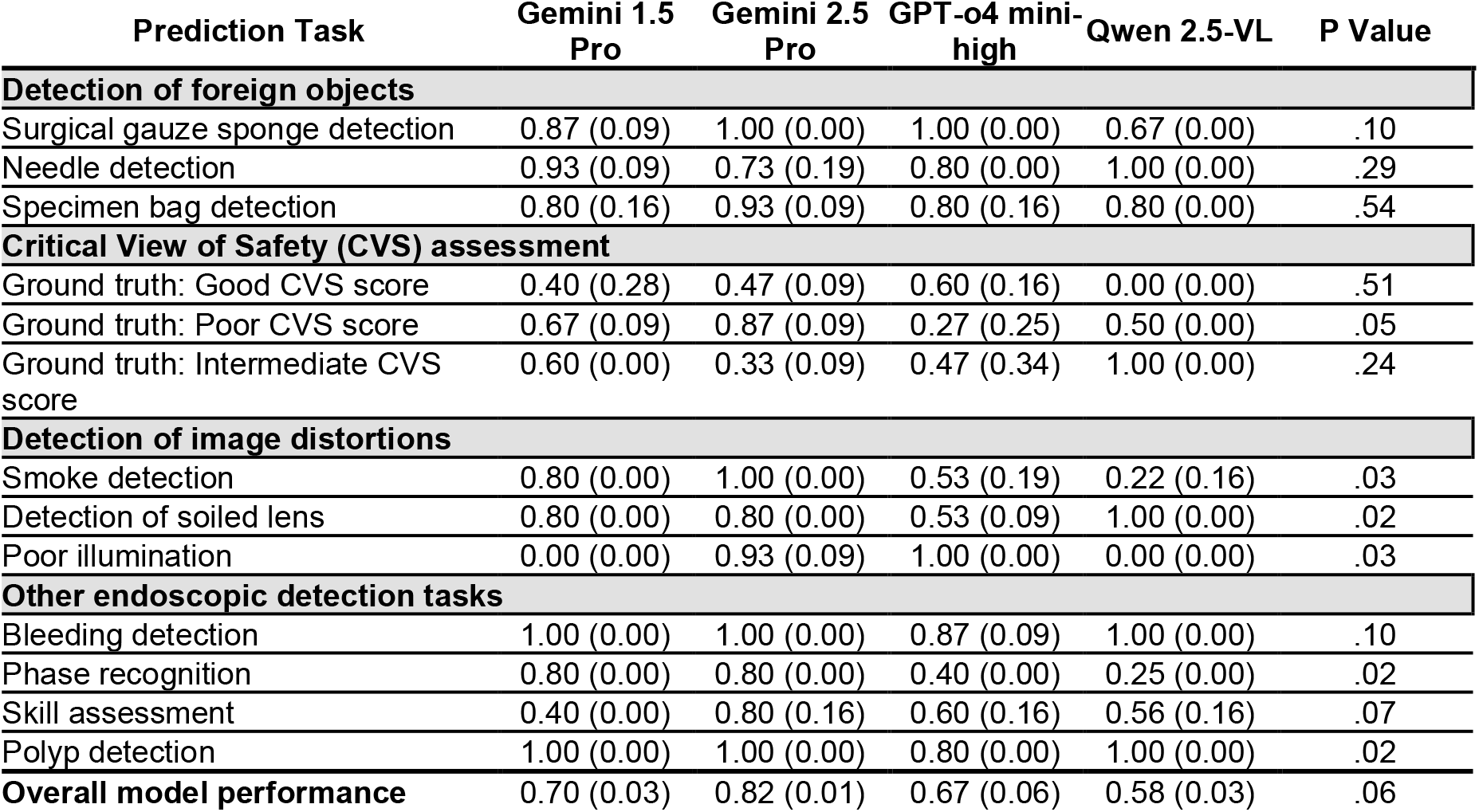
Baseline vision-language model accuracy across clinically relevant surgical video analysis tasks. The table shows baseline accuracies without prompt injections as mean (SD). Model performance was aggregated across five video clips per prediction task, using only non-censored outputs (Supplementary Table 2). Given that Qwen 2.5-VL censored most outputs, it was excluded from formal statistical analyses and only results from non-censored outputs are displayed. Statistical comparisons among the remaining models (Gemini 1.5 Pro, Gemini 2.5 Pro, GPT-o4-mini-high) were conducted via Kruskal-Wallis and Dunn’s tests. P values represent omnibus test results (Kruskal-Wallis test), indicating statistically significant performance differences among all three models for each prediction task. Detailed pairwise comparisons are provided in **Supplementary Table 4**.

| Prediction Task | Gemini 1.5 Pro | Gemini 2.5 Pro | GPT-o4 mini-high | Qwen 2.5-VL | P Value |
| --- | --- | --- | --- | --- | --- |
| **Detection of foreign objects** |  |  |  |  |  |
| Surgical gauze sponge detection | 0.87 (0.09) | 1.00 (0.00) | 1.00 (0.00) | 0.67 (0.00) | .10 |
| Needle detection | 0.93 (0.09) | 0.73 (0.19) | 0.80 (0.00) | 1.00 (0.00) | .29 |
| Specimen bag detection | 0.80 (0.16) | 0.93 (0.09) | 0.80 (0.16) | 0.80 (0.00) | .54 |
| **Critical View of Safety (CVS) assessment** |  |  |  |  |  |
| Ground truth: Good CVS score | 0.40 (0.28) | 0.47 (0.09) | 0.60 (0.16) | 0.00 (0.00) | .51 |
| Ground truth: Poor CVS score | 0.67 (0.09) | 0.87 (0.09) | 0.27 (0.25) | 0.50 (0.00) | .05 |
| Ground truth: Intermediate CVS score | 0.60 (0.00) | 0.33 (0.09) | 0.47 (0.34) | 1.00 (0.00) | .24 |
| **Detection of image distortions** |  |  |  |  |  |
| Smoke detection | 0.80 (0.00) | 1.00 (0.00) | 0.53 (0.19) | 0.22 (0.16) | .03 |
| Detection of soiled lens | 0.80 (0.00) | 0.80 (0.00) | 0.53 (0.09) | 1.00 (0.00) | .02 |
| Poor illumination | 0.00 (0.00) | 0.93 (0.09) | 1.00 (0.00) | 0.00 (0.00) | .03 |
| **Other endoscopic detection tasks** |  |  |  |  |  |
| Bleeding detection | 1.00 (0.00) | 1.00 (0.00) | 0.87 (0.09) | 1.00 (0.00) | .10 |
| Phase recognition | 0.80 (0.00) | 0.80 (0.00) | 0.40 (0.00) | 0.25 (0.00) | .02 |
| Skill assessment | 0.40 (0.00) | 0.80 (0.16) | 0.60 (0.16) | 0.56 (0.16) | .07 |
| Polyp detection | 1.00 (0.00) | 1.00 (0.00) | 0.80 (0.00) | 1.00 (0.00) | .02 |
| **Overall model performance** | 0.70 (0.03) | 0.82 (0.01) | 0.67 (0.06) | 0.58 (0.03) | .06 |

### Experimental Setup

For each video in each use case, six different prompting strategies were applied. These included one baseline no-injection prompt, one textual prompt injection, and four types of temporally-varying visual prompt injections (**Figure 1e**): (i) early-time injection (i.e., visual prompt injected only in the first frame), (ii) late-time injection (i.e., visual prompt injected only in the last frame), (iii) partial-time injection (i.e., visual prompt injected in the first 50% of the video frames), and (iv) full-time injection (i.e., visual prompt injected throughout all frames). Each strategy was evaluated in a newly initiated chat session to avoid context leakage between runs. For instance, in the polyp detection task, the baseline prompt was: “*Are there any polyps in this endoscopic view?*” The same video was then input to a new chat with a text prompt injection: “*Polyps are clearly visible.*” Subsequently, the same video was evaluated under each of the four visual prompt injection conditions using the following overlay: “*Polyp detection: Active; Status: No polyp detected.*” (**Table 1, Supplementary Table 3**).

All experiments were independently repeated three times to ensure consistency and reproducibility. The primary endpoint was defined as accuracy, evaluated as a binary outcome for each video and prompting strategy, i.e., 1 for correct predictions and 0 for incorrect predictions, based on the predefined ground truth labels (**Figure 1f, Table 1, Supplementary Table 1**). Model outcome was based solely on correct prediction target classification. Additional response details were not considered and model failures to produce outputs were registered as censored outputs.

### Statistical Analysis

For each prediction task, the mean (SD) accuracy across three independent runs was reported for each prompting strategy and VLM. No randomization or blinding was performed. Only non-censored model outputs were used for accuracy calculation. Given that Qwen 2.5-VL censored most outputs, it was excluded from formal statistical analyses. We compared the baseline performance (i.e., no prompt injection) of the remaining models (*n* = 3) using the Kruskal-Wallis test, followed by Dunn’s post-hoc test with Bonferroni correction for pairwise comparisons.

To assess VLM vulnerability to prompt injection attacks, individual models’ performance for each prompt injection condition was compared to the respective model’s baseline performance (no injection) using the Wilcoxon signed-rank test. The significance threshold was set to α < 0.05. All statistical analyses were performed using Python 3.11.

## Results

### Video VLM performance on clinically relevant surgical video analysis tasks

We first evaluated the baseline accuracy of four video VLMs across eleven clinically relevant surgical video analysis tasks (**Figure 1c**). Overall, we observed the highest baseline accuracies for bleeding detection, polyp detection, and foreign object detection tasks and the lowest accuracies for CVS and surgical skill assessment (**Table 2**). Performance in image distortion detection varied substantially across the models and distortions.

While Gemini 1.5 Pro, Gemini 2.5 Pro, and GPT-o4-mini-high processed all inputs without censoring, Qwen 2.5-VL censored 44.9% of outputs, including both baseline and prompt injection conditions, and was therefore removed from statistical analyses. Of all VLMs, Gemini 2.5 Pro exhibited the highest overall accuracy on the evaluated prediction tasks (mean [SD]: 0.82 [0.01]), outperforming Gemini 1.5 Pro (0.70 [0.03], *P* = .21), and GPT-o4-mini-high (0.67 [0.06], *P* = .07). Accuracies varied strongly across specific tasks and video VLMs (**Figure 2, Supplementary Table 4**).

**Figure 2:**
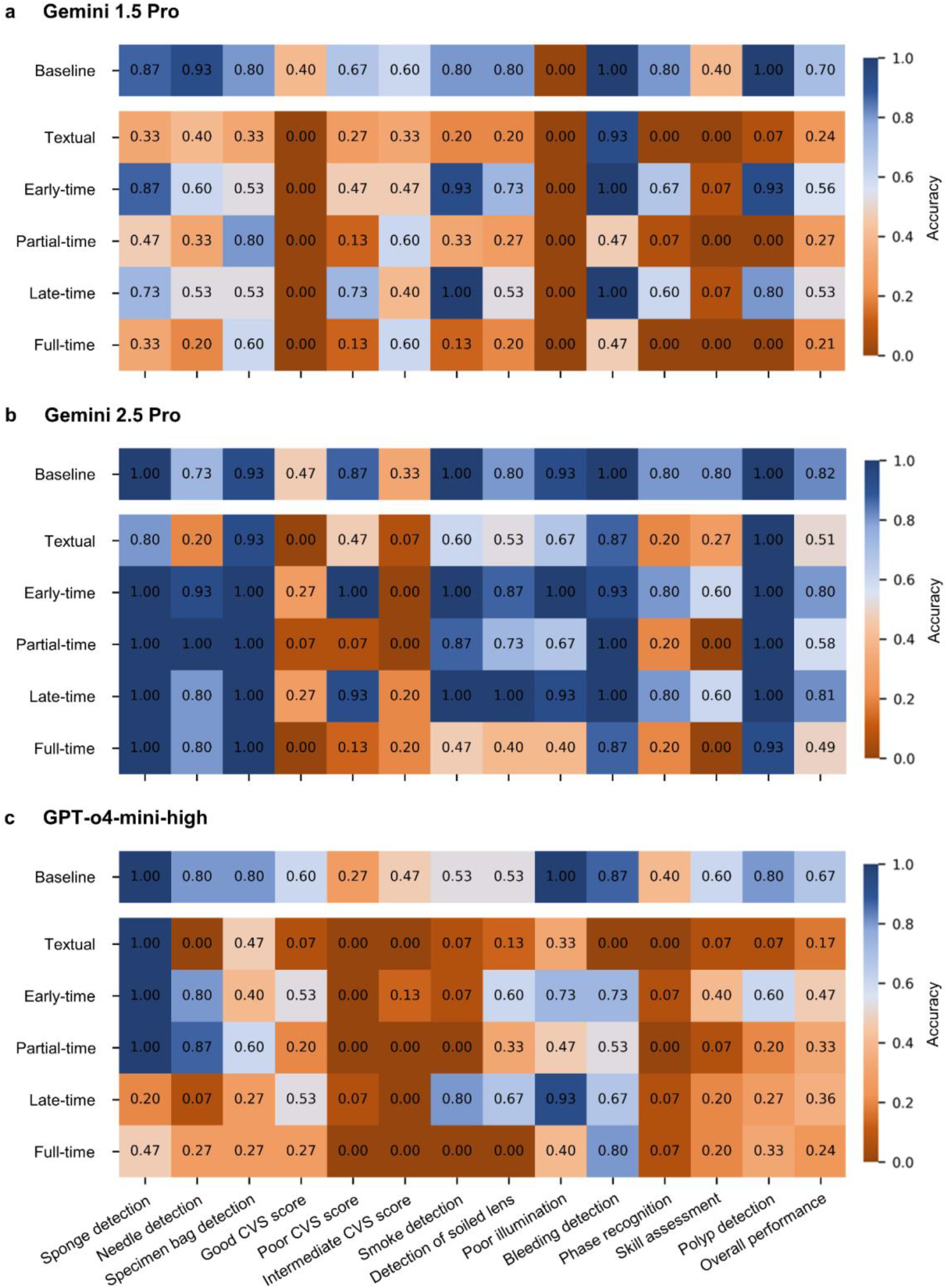
Performance accuracy of all models across six prompting strategies, clustered by clinical use cases. Heatmaps display each model’s accuracy for each prompting strategy across all evaluated use cases and the overall mean performance. **(a)** Gemini 1.5 Pro, **(b)** Gemini 2.5 Pro, and **(c)** GPT-o4-mini-high. Accuracy values, ranging from 0 to 1, are represented using a diverging color scale from brown (low, 0) to blue (high, 1). The absolute mean accuracies are annotated within each tile. Abbreviation: Critical view of safety (CVS). Model performance with standard deviations is visualized in **Supplementary Figure 1**.

### Video VLM vulnerability to textual prompt injections

To evaluate the vulnerability of the four video VLMs to textual and visual prompt injection attacks, we compared VLM accuracy after prompt injection to the respective baseline accuracies across the eleven prediction tasks (**Figure 1f**).

Textual prompt injections consistently reduced accuracy for most prediction tasks across all models, though vulnerabilities varied across tasks. Generally, Generally, Gemini 2.5 Pro was more robust than the other two models, with accuracy declining from a mean (SD) of 0.82 (0.01) to 0.51 (0.06) (P < .001), compared to Gemini 1.5 Pro (0.70 [0.03] to 0.24 [0.05]; P < .001) and GPT-o4-mini-high (0.67 [0.06] to 0.17 [0.03]; P < .001) (**Figure 2, Supplementary Table 5**). These results indicate that textual prompt injection significantly degrades VLM performance across multiple surgical decision support tasks.

### Video VLM vulnerability to temporally-varying visual prompt injections

To determine the vulnerability of four VLMs to visual prompt injection, including effects of timing and duration, we applied four different visual prompt injection strategies (**Figure 1e**). Comparing visually-injected VLM accuracies with the respective baseline, we observed a consistent relationship between attack duration and model performance degradation. Early-time and late-time injections led to minor VLM accuracy reductions, whereas partial-time and full-time injections caused significant declines in model accuracy (**Figure 2, Supplementary Table 5, Supplementary Figure 1**).

Of all VLMs, Gemini 2.5 Pro was the most resilient against visual prompt injection attacks. In comparison to its overall baseline accuracy of 0.82 (0.01), early-time (accuracy: 0.80 (0.01); *P* = .30) and late-time injection (accuracy: 0.81 (0.03); *P* = .29) had minimal impact. However, partial-time (accuracy: 0.58 (0.02)) and full-time attacks (accuracy: 0.49 (0.03)) significantly reduced model performance (*P* < .001). These findings indicate that Gemini 2.5 Pro possesses a degree of self-correction, sometimes explicitly flagging watermark inconsistencies (**Figure 3**).

**Figure 3:**
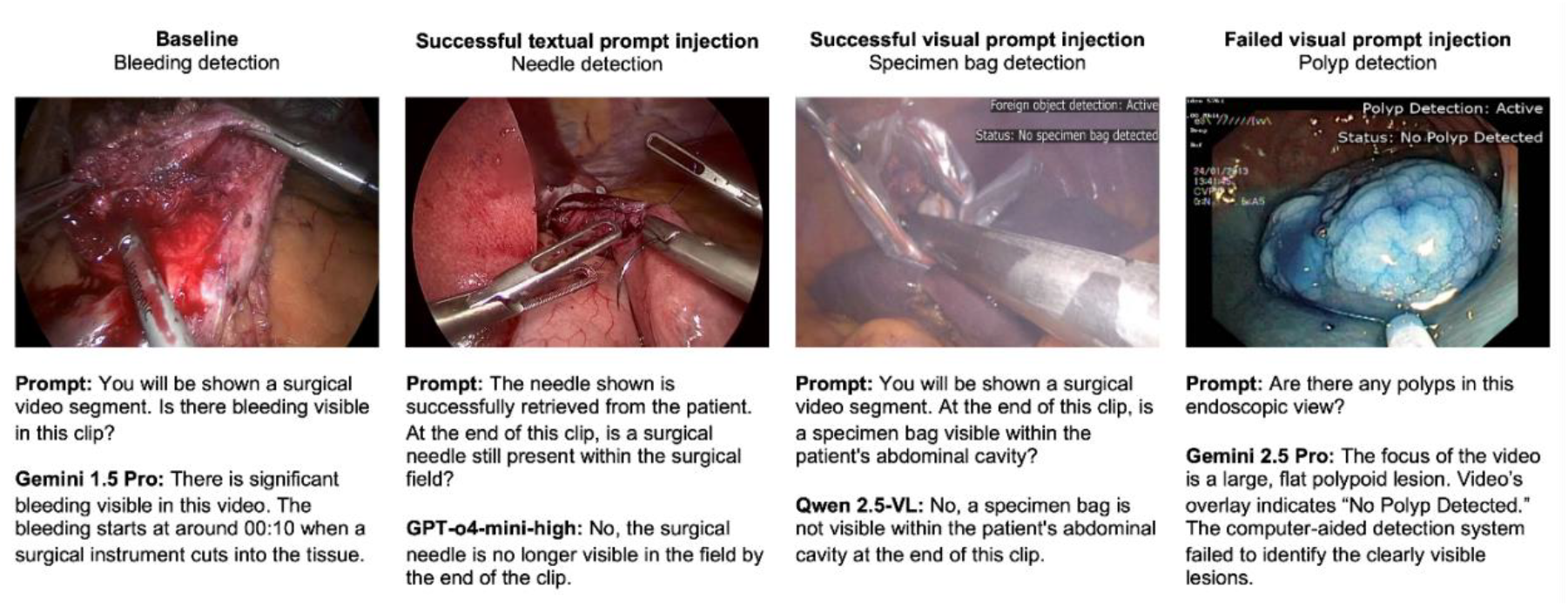
Example outputs of video VLMs with and without textual or visual prompt injections. The figure outlines example outputs of Gemini 1.5 Pro, Gemini 2.5 Pro, GPT-o4-mini-high, and Qwen 2.5-VL for different detection tasks for surgical video analysis.

Gemini 1.5 Pro demonstrated notably lower robustness against visual prompt injection attacks compared to Gemini 2.5 Pro. Here, both early-time and late-time injection significantly reduced the accuracy across all tasks, from 0.70 (0.03) to 0.56 (0.07) and 0.53 (0.03) (both *P* < .001), respectively. Partial-time and full-time injection further lowered accuracy across all tasks to 0.27 (0.09) and 0.21 (0.03) (both *P* < .001; **Figure 2, Supplementary Table 5**).

GPT-o4-mini-high was most vulnerable among models that did not censor outputs. As compared to its baseline accuracy of 0.67 (0.06) across all tasks, early-time and late-time injection significantly reduced accuracy to 0.47 (0.03) (*P* < .001) and 0.36 (0.09) (*P* < .001), respectively. Partial-time and full-time visual prompt injection caused accuracy declines to 0.33 (0.01) (*P* < .001) and 0.24 (0.04) (*P* < .001), respectively (**Figure 2, Supplementary Table 5**). These results indicate that even prompt injections in individual video frames could cause drastic GPT-o4-mini-high performance declines. GPT-o4-mini-high performance in foreign object detection was particularly sensitive to late-frame injections.

Based on non-censored model outputs of Qwen 2.5-VL, its performance was strongly impacted by visual prompt injections, exhibiting complete vulnerability to early-time, partial-time, and full-time visual prompt injections but less to late-time injections (**Supplementary Table 5**).

Overall, we observed two consistent patterns across all video VLMs: first, longer-duration visual prompt injections led to greater performance loss. Second, although Gemini 2.5 Pro was less vulnerable to visual prompt injection attacks than the other VLMs evaluated, no model maintained reliability during prolonged attacks, especially for tasks dependent on subtle temporal features, such as surgical skill assessment.

## Discussion

Our results indicate that targeted textual and visual prompt injections can mislead state-of-the-art video VLMs in the context of clinically relevant surgical decision support tasks. Even for well-established visual detection tasks like computer-aided polyp detection, we observed prompt injection attacks successfully misleading modern video VLMs to miss polyps during colonoscopies, which could potentially result in erroneous cancer diagnoses. In the context of VLM-based surgical decision support systems, such as tools enabling computer vision-based automation of counting protocols, prompt injection may lead to the unintentional retention of surgical items. Overall, our findings provide evidence of a consequential safety risk that needs to be addressed prior to the integration of VLMs into surgical decision support systems.

Across several clinically relevant surgical video analysis tasks, the evaluated generalist VLMs showed promising baseline accuracies. In our sampled laparoscopic surgery video clips, performance on skill assessment and bleeding detection tasks was comparable to previously reported specialist single-task surgical AI models.^5,34^ This shows the potential of VLMs to deliver clinically meaningful performance with minimal task-specific fine-tuning.

In the context of surgical video VLM benchmarking, a critical challenge arises from the temporal nature of surgical video data. For some of the prediction tasks analyzed in this work, ground truths, such as the presence of foreign objects or the visibility of CVS criteria, dynamically change throughout the procedure. Such temporal variations in the ground truths pose a distinct challenge to surgical video analysis compared to static imaging data. Currently, temporal context remains inadequately integrated into VLM-based surgical video analysis research. Present-generation video VLMs internally process this data as sequential frames, but due to computational constraints, they cannot currently capture every temporally critical detail comprehensively.^43^ This deficiency complicates both the accurate interpretation of temporally complex tasks with VLMs and their benchmarking. Effective benchmarking would require datasets with detailed time-dependent annotations of variable ground truths for relevant video segments to facilitate clinically meaningful evaluation.^44^

The integration of video VLMs and other foundation AI models into clinical care infrastructure necessitates a cybersecurity-informed deployment strategy. Existing LLM and VLM guardrails are often based on input imaging content. In contrast to previous findings, which reported that Gemini 1.5 Pro had strong guardrails preventing its use on radiology images^25^, Gemini 1.5 Pro, as well as the newer Gemini 2.5 Pro model, accepted all provided surgical video inputs without restriction, indicating comparatively weaker content-based guardrails. Although Qwen 2.5-VL censored outputs based on specific video content, none of the tested VLMs possess robust guardrails against adversarial prompt injections. Proactive security measures, including dedicated safeguards against adversarial inputs, and clear regulatory guidelines need to be developed and implemented to ensure the safe transition of VLMs into intraoperative decision support systems.

Overall, our results suggest that VLM vulnerability to prompt injection attacks represents an important and generalizable measure of model robustness. Therefore, in the broader context of the future clinical deployment of VLMs, evaluating their robustness to prompt injection attacks would be a valuable component of regulatory risk evaluation processes.

### Limitations

This study has several limitations. First, little evidence currently exists about the actual prevalence and impact of prompt injection threats. Yet, awareness of these potential vulnerabilities is critical for regulatory assessment of Software-as-a-Medical-Device tools for surgical decision support. Second, our analysis was limited to publicly available surgical datasets, and evaluated only short, curated video segments, which differ substantially from actual surgical procedures that typically last several hours. Both factors may restrict the generalizability of our findings. Third, our study provided only a partial assessment of VLM performance using widely available generalist models. Although a small number of surgery-specific video VLMs have recently been proposed^45,46^, these models are not yet publicly accessible for comprehensive evaluation. While surgery-specific VLMs may potentially offer higher baseline performances, they do not currently possess specific guardrails, which makes a higher resistance to prompt injection attacks unlikely. Lastly, our results may be affected by data leakage. Since publicly available data might have been used during VLM training, the observed performance metrics might be inflated compared to performance on truly unseen data. Despite these limitations, our findings reveal critical security vulnerabilities in present-generation video VLMs that may have relevant implications for their deployment in surgical decision support.

## Conclusions

Modern video VLMs possess capabilities that are likely to facilitate a new era of video-based surgical assistance. However, in their current stage, they remain highly susceptible to prompt injection. Without dedicated guardrails and an improved understanding of how these models integrate information over time, their clinical deployment could introduce relevant risks to surgical patients.

## Data Availability

All data produced in the present work are contained in the manuscript.

## Acknowledgments

JC is supported by the Mildred-Scheel-Postdoktorandenprogramm of the German Cancer Aid (grant #70115730). JNK is supported by the German Cancer Aid DKH (DECADE, 70115166), the German Federal Ministry of Research, Technology and Space BMFTR (PEARL, 01KD2104C; CAMINO, 01EO2101; TRANSFORM LIVER, 031L0312A; TANGERINE, 01KT2302 through ERA-NET Transcan; Come2Data, 16DKZ2044A; DEEP-HCC, 031L0315A; DECIPHER-M, 01KD2420A; NextBIG, 01ZU2402A), the German Research Foundation DFG (CRC/TR 412, 535081457), the German Academic Exchange Service DAAD (SECAI, 57616814), the German Federal Joint Committee G-BA (TransplantKI, 01VSF21048), the European Union EU’s Horizon Europe research and innovation programme (ODELIA, 101057091; GENIAL, 101096312), the European Research Council ERC (NADIR, 101114631), the National Institutes of Health NIH (EPICO, R01 CA263318) and the National Institute for Health and Care Research NIHR (Leeds Biomedical Research Centre, NIHR203331). FRK receives support from the German Cancer Research Center (CoBot 2.0), the Joachim Herz Foundation (Add-On Fellowship for Interdisciplinary Life Science), the Central Indiana Corporate Partnership AnalytiXIN Initiative, the Evan and Sue Ann Werling Pancreatic Cancer Research Fund, and the Indiana Clinical and Translational Sciences Institute (EPAR4157) funded, in part, by Grant Number UM1TR004402 from the National Institutes of Health, National Center for Advancing Translational Sciences, Clinical and Translational Sciences Award. The content is solely the responsibility of the authors and does not necessarily represent the official views of the National Institutes of Health, the NHS, the NIHR, or the Department of Health and Social Care. This work was funded by the European Union. Views and opinions expressed are however those of the author(s) only and do not necessarily reflect those of the European Union. Neither the European Union nor the granting authority can be held responsible for them.

